# Efficacy of a physical activity regimen on visual outcomes among persons newly diagnosed with abnormal glucose tolerance: Study protocol for a pilot randomized controlled trial

**DOI:** 10.1101/2024.04.10.24305644

**Authors:** Ebenezer Oduro Antiri, Thomas Hormenu, Edward Wilson Ansah, Stephen Ocansey, Rudolf Aaron Arthur, Eric Awlime-Ableh, Iddrisu Salifu, Benjamin Nyane, Augustine Mac-Hubert Gabla, Juliet Elikem Paku

## Abstract

Background: Abnormal Glucose Tolerance (AGT), which encompasses diabetes and prediabetes, is a growing health problem globally. It is affecting millions and predisposing such patients to several complications, including ocular complications. Physical activity has been found to improve glycemic levels, but the specific effect on visual outcomes in a newly diagnosed African population with AGT is yet to be explored fully. This pilot randomized controlled trial seeks to evaluate the efficacy of a physical activity intervention among newly diagnosed persons with AGT in Cape Coast, Ghana.

Methods: An ostensibly healthy population will be screened for AGT. Persons newly diagnosed with AGT will be recruited into the randomized controlled trial. About 50 newly diagnosed participants with AGT will undergo a 12-week moderate-to-vigorous physical activity (MVPA) intervention, specifically exercises that burn more than 3.0 metabolic equivalents (METs), to ascertain its effect on their visual acuity, contrast sensitivity, central visual field and diabetic retinopathy status. The participants will be randomized into a physical activity intervention group and a control group. Assessments will be done at baseline, and treatment outcomes will be done on the last day of the intervention for each participant.

Discussion: Physical activity is a proven lifestyle intervention that reduces glycemic levels in people with AGT. Unfortunately, many persons are unaware of their high level of glucose tolerance, which is associated with the deterioration of vision. This study intends to investigate and present findings on the advantages of physical activity interventions on visual outcomes among people with AGT. The study holds promise in informing evidence-based interventions for persons with AGT in poor economies.

## Background

Abnormal Glucose Tolerance (AGT), with all its negative health implication, is gradually becoming a regularized phenomenon with which society must contend. Instances of Impaired Glucose Tolerance (IGT), Impaired Fasting Glucose (IFG), and/or diabetes mellitus all identify as situations of AGT. The number of adults who suffer from only diabetes mellitus is estimated to experience a surge from 537 million adults to 783 million by 2045 (1). Additionally, there are also an estimated 541 million adults who have Impaired Glucose Tolerance (IGT) or prediabetes, making them highly susceptible to developing diabetes and its complications (1–3). Even without consideration of all other instances of Abnormal Glucose Tolerance (AGT), the estimated rate of growth of adults with diabetes mellitus is quite alarming.

In Ghana, the estimated prevalence of diabetes mellitus has been pegged at a high rate of 6.5% (4). As typical on the global scene, Type 2 Diabetes Mellitus (T2DM) is the most common in Ghana. It is important to acknowledge that instances of Abnormal Glucose Tolerance (AGT) such as overt diabetes have extensive precarious effects on an individual’s health system, such as one’s ocular health (5). Diabetic Retinopathy (DR), which is the one of most common microvascular complications of T2DM (6), has the tendency to cause total blindness if not managed properly (7). According to the literature (7), almost 60% of T2DM patients are found to have some form of diabetic retinopathy. Unfortunately, diabetic retinopathy has been a leading cause of adult-onset blindness in the past decade (8).

Type 2 Diabetes Mellitus (T2DM) is heavily influenced by both modifiable and partially adjustable risk factors, making it the most modifiable kind of diabetes (1,9). For example, previous studies have demonstrated that lifestyle changes, including a healthier diet and more physical activity significantly lower the risk of developing complications from T2DM (10–12). They also show that the vice versa is also possible where there can be complex complications from T2DM. It has been observed that there are numerous health advantages of physical activity such as improving glucose control, lipid profiling, and cardiovascular health, which help manage instances of AGT (13). Such demonstrations and observations suggest that in the instance of Diabetic Retinopathy (DR), for example, physical activity would have a direct impact on its management.

In fact, there have been studies that have investigated the associations between hyperglycemia (as associated with AGT) and visual functions, with these studies showing an observable reduction in visual functions due to hyperglycemia (14–20). Thus, these studies recommended lifestyle interventions like dietary changes and increased physical activity to improve visual outcomes. Further clinical trials also reported some positive change in visual functioning as a result of physical activity participation (21,22). However, many of the studies, as with the clinical trials, did not include an African population and persons with the prediabetes condition (5,21–23). Many of those studies rather focused on people diagnosed with diabetes who are likely to be on other prescribed interventions (5,21–23). Additionally, they assessed physical activity levels subjectively, by relying on patients’ self-reported data (21,22).

There is, therefore, the need for additional investigations into the related effects of physical activity and AGT, established through an objectively self-monitored physical activity regimen on visual outcomes in newly diagnosed persons with AGT from Africa. Understanding the direct impact of physical activity on AGT-related visual outcomes will enable the development of a more concise, evidence-based interventions for patients with AGT related ocular health issues. In that regard, the present study seeks interventions that are cost effective, person-specific, and target-driven. It has been suggested that cost-effective interventions are sustainable and can easily lead to actual lifestyle modification because they can be done without significant financial burden (24,25). Studies also show that such interventions need to be person-specific to be successful (26). For instance, persons with AGT may have varying characteristics, and tailoring physical activity interventions to their unique characteristics and needs is crucial for enhancing its adherence and effectiveness (27,28). Providing specific targets for a physical activity intervention is also another important factor, as it motivates participants and helps track their progress in adhering to the intervention (29).

In 2024, the International Diabetes Federation (IDF) and the International Agency for the Prevention of Blindness (IAPB) published a policy brief to help the development and implementation of measures to reduce diabetic eye disease worldwide (30). This trial protocol describes how the recommendations of developing contextually appropriate screening and treatment interventions, as well as building capacity in monitoring and evaluating progress of people with diabetic retinopathy can be implemented in an African population. Thus, the present study explores the effect of a cost-effective, person-specific, and target-driven physical activity regimen on visual outcomes among persons newly diagnosed with AGT in Cape Coast, Ghana.

## Objectives

The main objective of this study is to evaluate the feasibility of carrying out a double-blind Randomized Controlled Trial (RCT) and to investigate the effects of a moderate-to-vigorous physical activity (MVPA) regimen on visual outcomes among persons newly diagnosed with AGT in Cape Coast. The secondary objective is to investigate whether a physical activity regimen will influence the ability of the experimental group to cope with their daily visual demands, in comparison to the control group.

## Methods

### Study design

This study is a 12-week pilot and feasibility study with a double-blind, randomized control trial. The design will focus on the preliminary efficacy of physical activity to impact visual outcomes among newly diagnosed persons with AGT. The trial is designed based on the Standard Protocol Items: Recommendations for Interventional Trials (SPIRIT)(31) and will compare an experimental group (physical activity regimen) and an active placebo group (lifestyle change education), randomized into their respective groups in a 1:1 ratio. The trial design is further illustrated in Figures 1 and 2. The study protocol has been approved by the University of Cape Coast Institutional Review Board (UCCIRB) (UCCIRB/EXT/2022/27). The protocol is registered with the Pan African Clinical Trails Registry (PACTR202401462338744).

**Fig 1.**
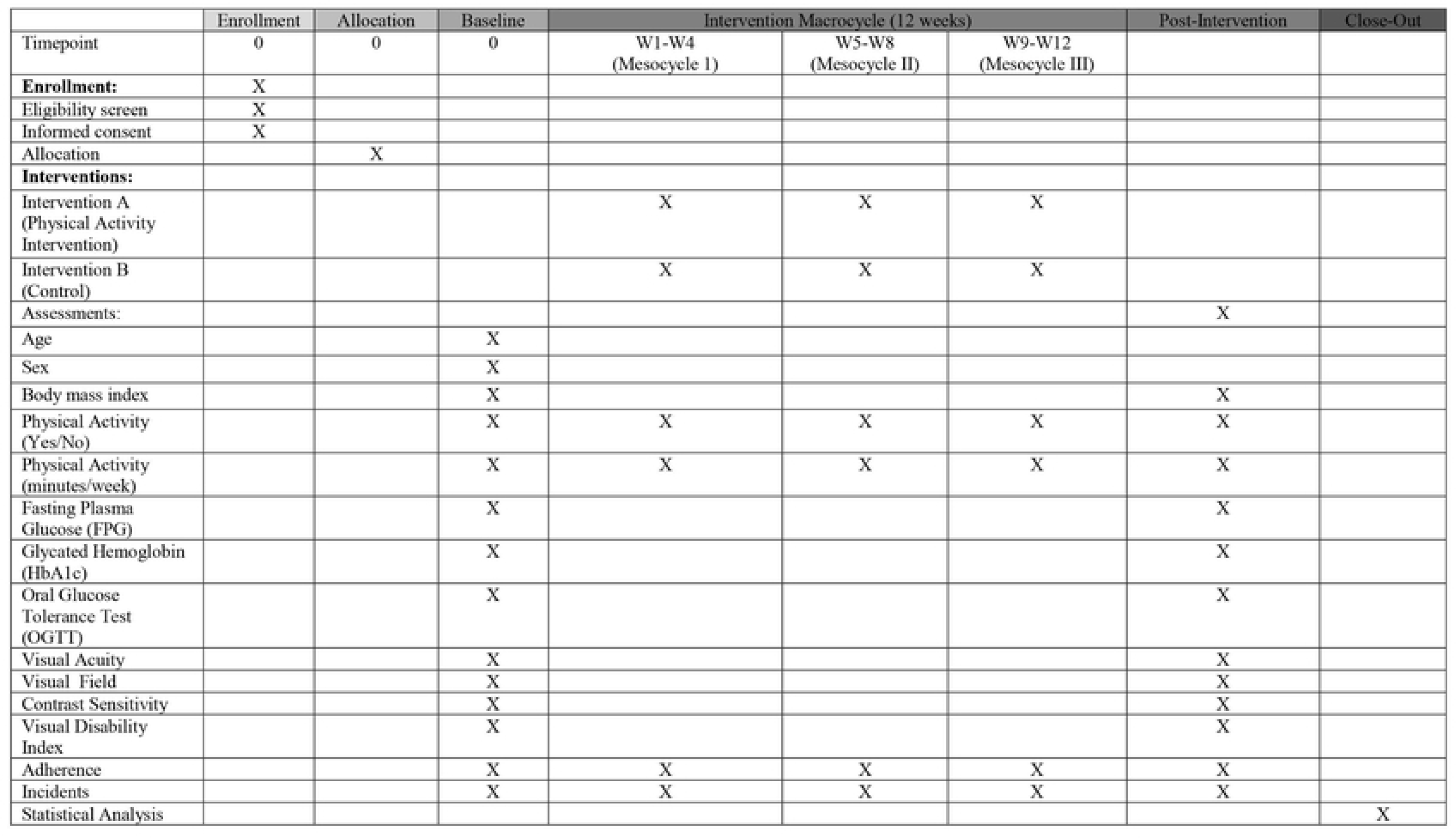
SPIRIT schedule of enrolment, interventions, and assessments

**Fig 2.**
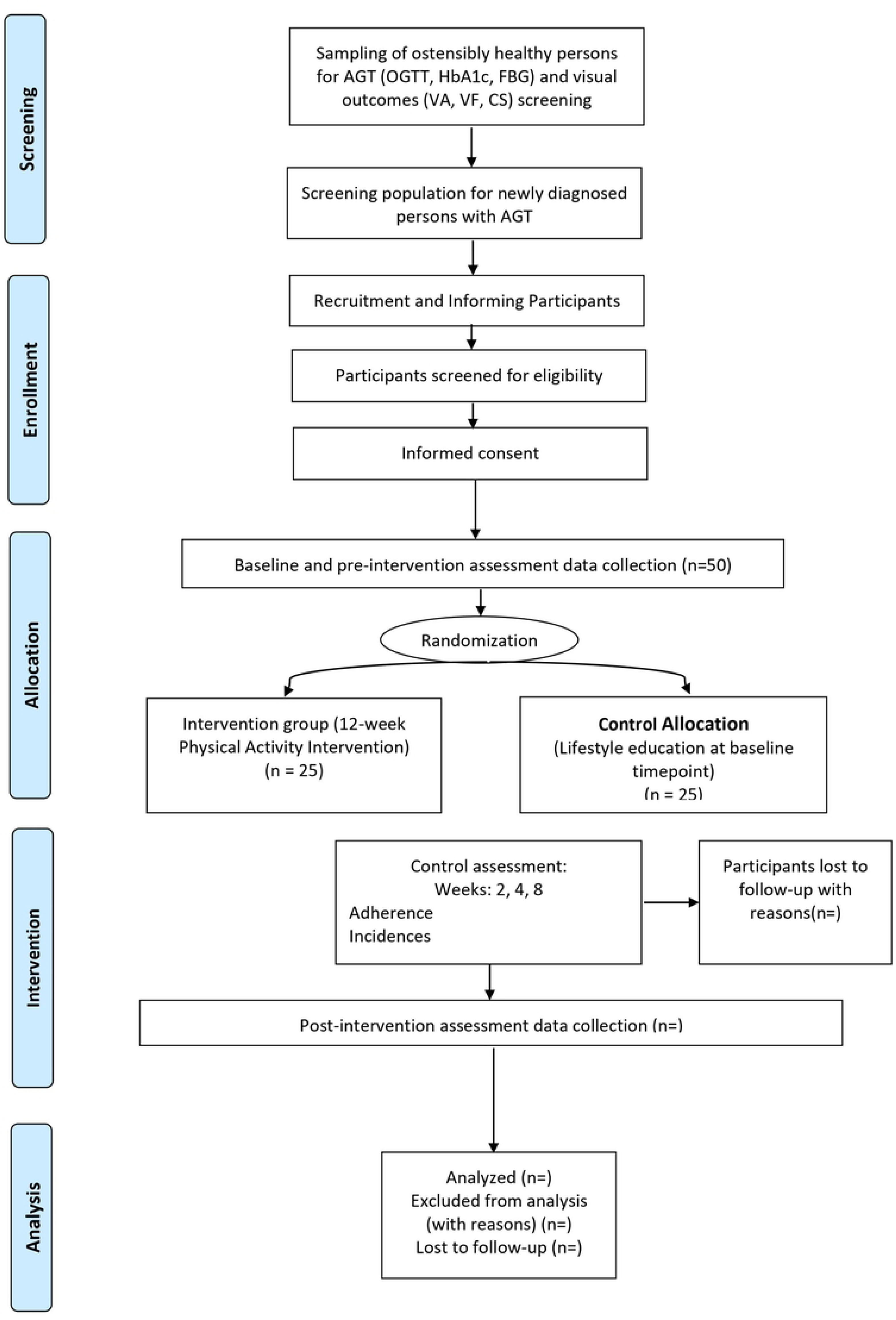
Flow chart of the RCT process for the participants in the study

### Settings and population

The study will be conducted at the Cardiometabolic Epidemiology Research Laboratory (CERL), University of Cape Coast. Recruitment will be carried out in communities in Cape Coast, Ghana. Moreover, the participants will be persons who have been newly diagnosed with AGT.

### Eligibilty criteria

All prospective participants in this trial will be well informed about the study. The participants will provide written or oral consent to participate in the trial. The inclusion criteria for the trial comprise: a) Newly diagnosed persons with AGT; and b) people between the ages of 25 and 70. The exclusion criteria for the trial comprise: a) Pregnant women and breastfeeding mothers; b) individuals with pre-existing diabetes (either type 1 or type 2); c) individuals with a BMI below 18.5 kg/m²; d) individuals with anemia; e) individuals with cardiac comorbidities or signs of cardiovascular instability; and f) individuals with any other contraindicated condition for moderate-intensity physical activity.

### Allocation and blinding

The participants in this study will be randomized into experimental or control groups, using the software www.randomizer.org. This process will be done by an independent researcher who will not take part in the intervention and evaluation stages of this trial. The randomization process will be done at a 1:1 ratio, with each participant given a special number.

Since the trial is double-blinded, both researchers and participants will be masked to the entire randomization process. As required, the researcher conducting the intervention will not take part in the evaluations while the evaluator will not know the allocations of the participants.

### Procedure

There will be a preliminary screening of the population to diagnose and identify individuals with AGT suffering from visual problems in the in the communities. This screening of the population in Coast to identify people eligible for the trial began in the first quarter of 2023 and ended in the final quarter of 2023. Individuals will be included in the clinical trial if they are diagnosed with AGT, being either prediabetes or diabetes for any of the Fasting Blood Glucose (FBG), Glycated Hemoglobin (HbA1c) and Oral Glucose Tolerance Test (OGTT) diagnostic tests. In addition, these individuals must have visual functioning deficits, including visual acuity, contrast sensitivity, and central visual field. Such individuals will then be informed of the purpose of the trial and their voluntary participation in the study will be solicited. Also, the Visual Disability Index of each participant will be measured using the Visual Disability Questionnaire (32). Those who will agree to take part in the study will then be given an informed consent form to sign (to take part in the baseline and pre-intervention assessments).

When the baseline and pre-intervention assessments have been conducted, an independent researcher will then assign the participants into either the experimental or control groups based on the 1:1 randomization using the randomization software. A researcher will be responsible for explaining the physical activity regimen to the experimental group, as well as setting up the fitness applications on their smartphones that will be used for estimating daily physical activity levels. Another researcher will also provide the necessary information to the control group for this trial.

The physical activity will be a macrocycle of 12 weeks, with three mesocycles consisting of four weeks each. Participants in the experimental group will be contacted every four weeks to enquire of the possible occurrences. This feedback will inform the next decision of whether the participants should move from one mesocycle to another, based on their adherence levels and willingness to continue. After the conclusion of the last week of the intervention phase, participants will be assessed at CERL on all parameters assessed at the baseline and pre-intervention stages.

### Assessments

The assessments for this trial will be conducted at CERL. All measures are expected to last around 120 minutes for each participant. The measures for this trial include:

*Fasting Blood Glucose (FBG)*: FBG will be measured after at least 8 hours of fasting using a glucometer. Blood samples will be taken from the participants and analyzed for blood glucose concentration, at mmol/L.

*Glycated Hemoglobin (HbA1c):* HbA1c will be measured from a blood sample that will be taken from participants using the HbA1c analyzer.

*Oral Glucose Tolerance Test (OGTT):* The glucose tolerance of the participants will be examined through the OGTT. Each participant will drink a 75g glucose solution after fasting overnight. Baseline glucose level will be measured, after which blood samples are taken at 1 hour and 2 hours after ingesting the glucose solution. The glucose levels then will be measured and analyzed. The glucose level recorded at exactly 2 hours after ingestion will determine whether the participant can be considered normal (less than 140 mg/dL), prediabetic (140 to 199 mg/dL), or diabetic (200 mg/dL or higher) (33).

*Body Mass Index:* Height and weight will be measured as means to gauge participants’ BMI. The participants will then be classified as severely underweight (less than 16.5kg/m^2^), underweight (under 18.5 kg/m^2^), normal weight (from 18.5 to 24.9 kg/m^2^), overweight (25 to 29.9 kg/m^2^), obesity (greater than or equal to 30 kg/m^2^), with sub-classifications of obesity class I (30 to 34.9 kg/m^2^), obesity class II (35 to 39.9 kg/m^2^) and obesity class III (greater than or equal to 40 kg/m^2^) (34).

*Visual Acuity (VA):* VA will be measured using the Early Treatment Diabetic Retinopathy Study (ETDRS) Chart for high contrast acuity. This chart reports VA on a log of the minimum angle of resolution (logMAR).

*Contrast Sensitivity (CS):* CS will be measured using a Pelli-Robson chart under standardized light conditions. This test measures the ability of the participants to discern letters at decreasing contrast levels.

*Central Visual Field (CVF):* CVF will be examined using the Amsler Chart. CVF is within 10 degrees radius from fixation and provides information on central vision function of the participants.

*Visual Disability Index (VDI):* Using the Visual Disability Questionnaire (VDQ), the VDI of the participants will be assessed to quantify the effects of visual impairment on an individual’s capacity to execute daily activities.

*Adherence:* Based on a similar clinical trial (35), adherence to the physical activity intervention will be defined and assessed as having done MVPA with the physical activity tracker on for more than 75% of the intervention period and having read more than 50% of program content.

*Incidents:* Incidents will be defined as any adverse, uncomfortable, or unexpected events that will jeopardize the ability and willingness of the participant to continue the clinical trial. This assessment will be case dependent, ranging from telephonic evaluation to clinical evaluation at CERL.

### Intervention

According to the key guidelines, adults with chronic conditions or disabilities, who are able, should do at least 150 minutes (2 hours and 30 minutes) to 300 minutes (5 hours) a week of moderate-intensity physical activity, or 75 minutes (1 hour and 15 minutes) to 150 minutes (2 hours and 30 minutes) a week of vigorous-intensity aerobic physical activity, or an equivalent combination of moderate- and vigorous-intensity aerobic activity (36,37). The comparison is that 1 minute of vigorous-intensity activity is equivalent to 2 minutes of moderate-intensity activity. That is, the lower limit of vigorous-intensity physical activity (6.0 METs) is twice the lower limit of moderate-intensity activity (3.0 METs). Therefore, 75 minutes of vigorous-intensity activity a week is roughly equivalent to 150 minutes of moderate-intensity activity a week.

This is the basis for the development of the tailored physical activity regimen for each of the participants. The main moderate-intensity physical activity adopted for this study is brisk walking while that of vigorous-intensity physical activity is running. The design and implementation mirrored a similar physical activity study that assessed the impact of physical activity on abnormal glucose tolerance and visual outcomes (22). The intervention period (macrocycle) was divided into three mesocycles (four weeks each), with fixed increments with each subsequent mesocycle. For participants with T2DM, physical activity should not be performed after taking oral hypoglycemic medication when they have not eaten.

### Intervention group

The participants in the experimental group will undergo moderate-intensity physical activity (150 minutes and above) or vigorous-intensity physical activity (75 minutes and above), or combine the two in an equivalent way. This will depend on the age, gender, and daily activity of the participants. Subsequently, the amount of time for physical activity will be increased by 75 minutes for moderate-intensity physical activity and 37.5 minutes for vigorous-intensity physical activity for every four weeks during the intervention phase. The physical activity will be structured, planned, individualized, and tracked during the period. The activity can be performed at home, work, or at fitness facilities. There will be no more than two consecutive days without undertaking the exercise.

The amount of time of physical activity will be self-tracked using the accelerometer and GPS sensors of the participants’ smartphones. Mobile phone fitness and health applications will be used to document the patient’s physical activity history. The Apple Health App will be used by participants who use IOS run mobile phones while the Google Fit App will be used by android phones users (in the cases where the brands of the android mobile phone does not have in-built fitness and health applications).

### Control group

Participants in the control group will receive health education, including materials focusing on the importance of regular physical activity for overall health and well-being. The education will cover general recommendations for physical activity, potential benefits, and strategies for incorporating movement into daily life. Unlike the intervention group, there will be no prescribed duration, intensity, or frequency of engaging in physical activity. The goal is to promote a general understanding of the benefits of physical activity without providing a specific, planned activity regimen. As opposed to the robust tracking system set up for the physical activities of the experimental group, the control group participants will be asked to keep a simple log or diary of their physical activities.

### Outcome Measures

#### Primary outcome: Visual outcomes

Adherence to a physical activity intervention would significantly improve visual outcomes in an African population with AGT. This outcome presupposes a difference in visual outcomes before and after the physical activity regimen has been completed by the participants. The visual outcomes being assessed in this trial will be visual acuity, contrast sensitivity, and central visual field of the participants, before and after the introduction of the physical activity intervention. Visual acuity will be assessed using the Early Treatment Diabetic Retinopathy Study (ETDRS) chart, contrast sensitivity will be assessed using the Pelli-Robson contrast sensitivity chart, and the central visual field (CVF) will be assessed using the Amsler Chart.

#### Secondary outcomes: Glycemic level, Visual Disability Index, Feasibility

##### Glycemic level

Adherence to a physical activity intervention significantly reduces glycemic levels in an African population with AGT. This outcome presupposes a difference in glycemic levels before and after the physical activity regimen has been completed by the participants. Glycemic levels in this trial will be assessed in three ways: Oral Glucose Tolerance Test (OGTT), Glycated Hemoglobin (HbA1c), and Fasting Blood Glucose (FBG).

##### Visual Disability Index (VDI)

The visual disability index involves a quantification of the effects of visual impairment on an individual’s capacity to execute daily activities. It offers a comprehensive evaluation of the effects of ocular conditions on the general functioning of a person. Visual Disability Index (VDI) will be assessed using the Visual Disability Questionnaire (VDQ) adapted from Shrestha and Kaiti (2014) who used the inventory to study diabetic patients.

### Feasibilty

Feasibility will be assessed by the number of people who are recruited for the trial and how well they adhere to the requirements of the trial. In addition, occurrences (positive or negative) will be assessed as well (38,39). This will be documented along with the clinical outcomes.

The clinical outcomes (FPG, HbA1c, OGTT, VA, CS, CVF) will be assessed at pre-intervention, after each mesocycle and post intervention. VDI will be measured only at pre-intervention and post-intervention. Concerning the feasibility outcome measures, data on incidence and adherence will be collected weekly, with recruitment already assessed at the pre-intervention stage.

### Sample size

A total of 50 participants are expected to be recruited for this study. There will be 20 participants in each group. An additional 10 will be recruited to cater for attrition or participant dropout as recommended for RCT pilot studies (40,41).

### Data management

A comprehensive database will be created and used to monitor the progress of each participant, including all assessments. All collected data will be strictly recorded and stored on a password-protected computer, accessible only to the research team, but kept with the principal investigator. An independent researcher will oversee the data collection process and ensures its safety. The analysis of the data will begin when the complete data for each assessment has been collected at all stages of the trial.

### Statistical analysis

SPSS software version 27.0 (SPSS Inc, Chicago, IL) will be used to manage and analyse the data. The normality of the data will be tested using the Shapiro-Wilk test. The significance level for all the statistical tests will be 0.05. Descriptive statistics will be done to provide an overview of the data. This would include frequencies for categorical variables, measures of central tendency and spread for continuous variables.

The difference in the primary outcome (visual outcomes improving after the physical activity regimen) will be analyzed using the mixed-design Analysis of Variance (ANOVA), with a within-subjects factor (pretest and posttest) and a between-subject factor (experimental and control) at p < 0.05. This is similar to a previous experimental study carried out among patients with diabetic retinopathy (22).

The secondary outcome or difference in glycemic levels after the physical activity intervention will be analyzed using a paired t-test to compare the changes in the visual disability index within each group from baseline to post-intervention. An independent-samples t-test will be used to assess the difference in glycemic levels between the intervention and control groups at post-intervention. This is similar to a study which investigated the effect of a physical activity promotion program on the adherence to physical activity among people living with type 2 diabetes (42).

The other secondary outcome of visual disability index, reducing after the physical activity intervention, will be assessed with within-group comparisons using a paired t-test to compare the changes in the visual disability index within each group, from baseline to post-intervention. Also, between-group comparisons will be made using independent-samples t-test to assess the changes in VDI between the experimental group and control group at post-intervention, with p-value less than 0.05 considered significant.

### Ethics

This study will adhere to the tenets of the Declaration of Helsinki on research involving human subjects (43). The protocol has also been approved by the UCCIRB (UCCIRB/EXT/2022/27). In addition, this research is registered with the Pan African Clinical Trails Registry - PACTR202401462338744.

Furthermore, the dataset from this study will only be accessible to the researcher responsible for analyzing the data. To maintain confidentiality and privacy of the participants, the data will be ciphered by assigning codes to conceal any details that might identify a participant.

## Discussion

Evidence suggests that before visible fundus changes appear in patients with abnormal glucose tolerance, there is some substantial inner retinal dysfunction caused by the elevated glycemic levels (17). This is characterized by morphological changes in the Retinal Ganglion Cell (RGC) and inner retinal layers of the patients (44). Accordingly, the condition could be caused by chronic exposure to high glucose levels that induces oxidative stress and activates inflammatory pathways in the retina, which in turn affects perception of vision and proper visual functioning (17,45,46).

There is a strong scientific evidence to suggest that non-pharmacological lifestyle changes, like regular physical activity, can reduce the risk of cardiovascular diseases (CVD), type 2 diabetes, metabolic syndrome, obesity, and certain types of cancer (36). Regular physical activity includes structured exercise in a variety of forms and it offers a net benefit for most individuals living with T2DM (7,11,35,47). For instance, regular physical activity enhances insulin sensitivity, increases cardiorespiratory fitness, improves glycemic control, reduces the risk of cardiovascular mortality, and enhances psychosocial well-being (13). The term “trained eye”, akin to the phrase “trained heart” used in cardiology, highlights the positive effects of physical activity on not only the health of the eye, but also on the eye and the visual system (48).

Many studies have investigated the effect of non-pharmacological interventions on selected visual functions, and recorded some positive change in visual functioning (21,22). An interventional study assessed the effect of a multicomponent nutritional formula on visual functions among people with both short and long-term diabetes and found that the intervention offered some overall protection of visual function associated with the onset and progression of DR in both groups (21). However, this study was conducted at a single center and did not discover any significant intergroup difference. To improve on the previous results, another study (22) designed an individual-specific tailored physical activity intervention to improve compliance and elicit uniform results, regardless of the varying glycemic levels among the participants. The study recorded decreased glycemic levels and a resulting improvement in visual functioning, especially visual acuity. However, their did not include people with prediabetes as the focus was on diabetic patients who are likely to be on other interventions (22). Thus, it is important to also assess the efficacy of these interventions on both diabetic and prediabetic persons. Also, adherence in these studies was self-reported by the patients, making such measurements very subjective. Therefore, it is important to assess the efficacy of a physical activity regimen on visual outcomes among newly diagnosed patients with AGT, and that adherence to the regimen is measured objectively.

This trial has the element of objective tracking of the physical activity regimen. Subjective tracking where patients record how much physical activity they have done poses a challenge because of the case of self-desirability bias it presents (49). There is a growing emphasis on self-monitoring applications that allow patients to measure and track their own physical health parameters (weight, physical activity, blood pressure, blood glucose level, lung function), and where patients’ willingness to self-monitor might be associated with disease controllability (50). In fact, among patients with diabetes mellitus, asthma, hypertension, rheumatism and migraine, those with diabetes mellitus have been found to be more willing to self-monitor their health parameters (50).

At the conclusion of this trial, we expect to identify changes in visual outcomes, especially visual acuity, as was found in a recent study that tested for similar outcomes (22). Also, it is expected that there will be changes in the VDI of the participants as has been observed in a previous study (32). Generally, this trial seeks to confirm the associations that have been comprehensively described by a recent systematic review on the topic area (23).

## Strengths and Weaknesses

While there will be efforts to ensure blinding, there remains a possibility of bias because the experimental group will be motivated more than the control group to be more physically active. The other limitation is that the study is based on participants following the prescribed exercise regimen to the latter. This may vary due to personal preferences, lifestyle, or challenges to exercise.

Despite these limitations, the study’s strengths lie in its rigorous design, detailed outcome measures, objectives, and accurate adherence monitoring. While adhering to the ethical guidelines, this research intends to give a comprehensive analysis of the feasibility and preliminary efficacy of physical activity interventions for improving visual outcomes among people with AGT.

## Data Availability

No datasets were generated or analysed during the current study. All relevant data from this study will be made available upon study completion.

## Clinical trial registration

This trial has been registered with the Pan African Clinical Trails Registry with the identifier: PACTR202401462338744.

## Supporting Information

Supporting Information file S1.

Supporting Information file S2.

## References

1. International Diabetes Federation. IDF Diabetes Atlas. 10th editi. Brussels; 2021.

2. Bartnik M, Malmberg K, Hamsten A, Efendic S, Norhammar A, Silveira A, et al. Abnormal glucose tolerance--a common risk factor in patients with acute myocardial infarction in comparison with population-based controls. J Intern Med. 2004 Oct;256(4):288–97.

3. Palmert MR, Gordon CM, Kartashov AI, Legro RS, Emans SJ, Dunaif A. Screening for Abnormal Glucose Tolerance in Adolescents with Polycystic Ovary Syndrome. J Clin Endocrinol Metab [Internet]. 2002 Mar 1;87(3):1017–23. Available from: 10.1210/jcem.87.3.8305

4. Asamoah-Boaheng M, Sarfo-Kantanka O, Tuffour A, Eghan B, Mbanya J. Prevalence and risk factors for diabetes mellitus among adults in Ghana: a systematic review and meta-analysis. Int Health. 2019;11(2):83–92.

5. Faiq MA, Sengupta T, Nath M, Velpandian T, Saluja D, Dada R, et al. Ocular manifestations of central insulin resistance. Neural Regen Res. 2023 May;18(5):1139–46.

6. Mansour A, Mousa M, Abdelmannan D, Tay G, Hassoun A, Alsafar H. Microvascular and macrovascular complications of type 2 diabetes mellitus: Exome wide association analyses. Front Endocrinol (Lausanne) [Internet]. 2023;14:1143067. Available from: https://www.frontiersin.org/articles/10.3389/fendo.2023.1143067

7. Bukht M, Ahmed K, Hossain S, Masud P, Sultana S, Khanam R. Association between physical activity and diabetic complications among Bangladeshi type 2 diabetic patients. Diabetol Metab Syndr. 2019;13(1):806–9.

8. Bourne RRA, Stevens GA, White RA, Smith JL, Flaxman SR, Price H, et al. Causes of vision loss worldwide, 1990-2010: A systematic analysis. Lancet Glob Heal. 2013 Dec;1(6):e339–49.

9. Issaka A, Paradies Y, Stevenson C. Modifiable and emerging risk factors for type 2 diabetes in Africa: a systematic review and meta-analysis protocol. Syst Rev. 2018;7:139.

10. Davies MJ, Aroda VR, Collins BS, Gabbay RA, Green J, Maruthur NM, et al. Management of hyperglycaemia in type 2 diabetes, 2022. A consensus report by the American Diabetes Association (ADA) and the European Association for the Study of Diabetes (EASD). Diabetologia. 2022;65(12):1925–66.

11. Denche-Zamorano Á, Mendoza-Muñoz DM, Barrios-Fernandez S, Perez-Corraliza C, Franco-García JM, Carlos-Vivas J, et al. Physical activity reduces the risk of developing diabetes and diabetes medication use. Healthcare. 2022 Dec;10(12):2479.

12. Tuomilehto J, Lindström J, Eriksson JG, Valle TT, Hämäläinen H, Ilanne-Parikka P, et al. Prevention of type 2 diabetes mellitus by changes in lifestyle among subjects with impaired glucose tolerance. N Engl J Med. 2001 May;344(18):1343–50.

13. Carbone S, Del Buono MG, Ozemek C, Lavie CJ. Obesity, risk of diabetes and role of physical activity, exercise training and cardiorespiratory fitness. Prog Cardiovasc Dis. 2019;62(4):327–33.

14. Bao YK, Yan Y, Gordon M, McGill JB, Kass M, Rajagopal R. Visual field loss in patients with diabetes in the absence of clinically-detectable vascular retinopathy in a nationally representative survey. Invest Ophthalmol Vis Sci. 2019 Nov;60(14):4711– 6.

15. Gualtieri M, Bandeira M, Hamer RD, Damico FM, Moura ALA, Ventura DF. Contrast sensitivity mediated by inferred magno- and parvocellular pathways in type 2 diabetics with and without nonproliferative retinopathy. Invest Ophthalmol Vis Sci. 2011 Feb;52(2):1151–5.

16. Mirković J, Risimić D. Contrast sensitivity in patients with diabetes mellitus. Med Podml. 2018;69(4):35–9.

17. Nagai N, Mushiga Y, Ozawa Y. Evaluating fine changes in visual function of diabetic eyes using spatial-sweep steady-state pattern electroretinography. Sci Rep [Internet]. 2023;13(1):13686. Available from: 10.1038/s41598-023-40686-5

18. Pinilla I, Ferreras A, Idoipe M, Sanchez-Cano AI, Perez-Garcia D, Herrera LX, et al. Changes in frequency-doubling perimetry in patients with type I diabetes prior to retinopathy. Biomed Res Int. 2013;2013:341269.

19. Siersma V, Køster-Rasmussen R, Bruun C, Olivarius N de F, Brunes A. Visual impairment and mortality in patients with type 2 diabetes. BMJ open diabetes Res care. 2019;7(1):e000638.

20. Stavrou EP, Wood JM. Letter contrast sensitivity changes in early diabetic retinopathy. Clin Exp Optom. 2003 May;86(3):152–6.

21. Chous AP, Richer SP, Gerson JD, Kowluru RA. The Diabetes Visual Function Supplement Study (DiVFuSS). Br J Ophthalmol [Internet]. 2016 Feb 1;100(2):227–34. Available from: http://bjo.bmj.com/content/100/2/227.abstract

22. Pardhan S, Upadhyaya T, Smith L, Sharma T, Tuladhar S, Adhikari B, et al. Individual patient-centered target-driven intervention to improve clinical outcomes of diabetes, health literacy, and self-care practices in Nepal: A randomized controlled trial. Front Endocrinol (Lausanne). 2023;14:1076253.

23. AlQabandi Y, Nandula SA, Boddepalli CS, Gutlapalli SD, Lavu VK, Abdelwahab Mohamed Abdelwahab R, et al. Physical activity status and diabetic retinopathy: A review. Cureus. 2022 Aug;14(8):e28238.

24. Sathish T, Oldenburg B, Thankappan KR, Absetz P, Shaw JE, Tapp RJ, et al. Cost-effectiveness of a lifestyle intervention in high-risk individuals for diabetes in a low- and middle-income setting: Trial-based analysis of the Kerala Diabetes Prevention Program. BMC Med. 2020 Sep;18(1):251.

25. Singh K, Chandrasekaran AM, Bhaumik S, Chattopadhyay K, Gamage AU, Silva P De, et al. Cost-effectiveness of interventions to control cardiovascular diseases and diabetes mellitus in South Asia: A systematic review. BMJ Open. 2018;8(4):e017809.

26. Edlind M, Mitra N, Grande D, Barg FK, Carter T, Turr L, et al. Why effective interventions do not work for all patients: exploring variation in response to a chronic disease management intervention. Med Care. 2018 Aug;56(8):719–26.

27. Skoglund G, Nilsson BB, Olsen CF, Bergland A, Hilde G. Facilitators and barriers for lifestyle change in people with prediabetes: a meta-synthesis of qualitative studies. BMC Public Health [Internet]. 2022;22(1):553. Available from: 10.1186/s12889-022-12885-8

28. Ma JK, Floegel TA, Li LC, Leese J, De Vera MA, Beauchamp MR, et al. Tailored physical activity behavior change interventions: challenges and opportunities. Transl Behav Med. 2021 Dec;11(12):2174–81.

29. Collado-Mateo D, Lavín-Pérez AM, Peñacoba C, Del Coso J, Leyton-Román M, Luque-Casado A, et al. Key factors associated with adherence to physical exercise in patients with chronic diseases and older adults: An umbrella review. Int J Environ Res Public Health. 2021 Feb;18(4).

30. IAPB, IDF. Diabetic Retinopathy: A Call for Global Action. 2024.

31. Chan AW, Tetzlaff JM, Altman DG, Laupacis A, Gøtzsche PC, Krleža-Jerić K, et al. SPIRIT 2013 statement: defining standard protocol items for clinical trials. Ann Intern Med. 2013 Feb;158(3):200–7.

32. Shrestha GS, Kaiti R. Visual functions and disability in diabetic retinopathy patients. J Optom [Internet]. 2014;7(1):37–43. Available from: https://www.sciencedirect.com/science/article/pii/S188842961300023X

33. ElSayed NA, Aleppo G, Aroda VR, Bannuru RR, Brown FM, Bruemmer D, et al. Classification and diagnosis of diabetes: Standards of care in diabetes—2023. Diabetes Care [Internet]. 2022 Dec 12;46(Supplement_1):S19–40. Available from: 10.2337/dc23-S002

34. Weir CB, Jan A. BMI Classification Percentile And Cut Off Points. In Treasure Island (FL); 2024.

35. Kooiman TJM, de Groot M, Hoogenberg K, Krijnen WP, van der Schans CP, Kooy A. Self-tracking of physical activity in people with type 2 diabetes: A randomized controlled trial. Comput Inform Nurs. 2018 Jul;36(7):340–9.

36. Piercy KL, Troiano RP, Ballard RM, Carlson SA, Fulton JE, Galuska DA, et al. The physical activity guidelines for Americans. JAMA. 2018 Nov;320(19):2020–8.

37. DiPietro L, Buchner DM, Marquez DX, Pate RR, Pescatello LS, Whitt-Glover MC. New scientific basis for the 2018 U.S. Physical Activity Guidelines. J Sport Heal Sci. 2019 May;8(3):197–200.

38. Morris NS, Rosenbloom DA. CE: Defining and understanding pilot and other feasibility studies. Am J Nurs. 2017 Mar;117(3):38–45.

39. Resnick B. The definition, purpose and value of pilot research. Geriatr Nurs. 2015;36(2):S1–2.

40. Kieser M, Wassmer G. On the use of the upper confidence limit for the variance from a pilot sample for sample size determination. Biometrical J [Internet]. 1996 Jan 1;38(8):941–9. Available from: 10.1002/bimj.4710380806

41. Whitehead AL, Julious SA, Cooper CL, Campbell MJ. Estimating the sample size for a pilot randomised trial to minimise the overall trial sample size for the external pilot and main trial for a continuous outcome variable. Stat Methods Med Res. 2016 Jun;25(3):1057–73.

42. Eshete A, Mohammed S, Shine S, Eshetie Y, Assefa Y, Tadesse N. Effect of physical activity promotion program on adherence to physical exercise among patients with type II diabetes in North Shoa Zone Amhara region: a quasi-experimental study. BMC Public Health [Internet]. 2023;23(1):709. Available from: 10.1186/s12889-023-15642-7

43. World Medical Association. World Medical Association Declaration of Helsinki: Ethical principles for medical research involving human subjects. JAMA. 2013 Nov;310(20):2191–4.

44. Amato R, Catalani E, Dal Monte M, Cammalleri M, Cervia D, Casini G. Morpho-functional analysis of the early changes induced in retinal ganglion cells by the onset of diabetic retinopathy: The effects of a neuroprotective strategy. Pharmacol Res [Internet]. 2022;185:106516. Available from: https://www.sciencedirect.com/science/article/pii/S1043661822004625

45. Park HYL, Kim IT, Park CK. Early diabetic changes in the nerve fibre layer at the macula detected by spectral domain optical coherence tomography. Br J Ophthalmol. 2011 Sep;95(9):1223–8.

46. Spaide RF. Measurable aspects of the retinal neurovascular unit in diabetes, glaucoma, and controls. Am J Ophthalmol. 2019 Nov;207:395–409.

47. Amin M, Kerr D, Atiase Y, Yakub Y, Driscoll A. Understanding physical activity behavior in Ghanaian adults with type 2 diabetes: A qualitative descriptive study. J Funct Morphol Kinesiol. 2023;8(3):127.

48. Szalai I, Pálya F, Csorba A, Tóth M, Somfai G. The effect of physical exercise on the retina and choroid. Klin Monbl Augenheilkd. 2020;237(4):446–9.

49. Howitt C, Brage S, Hambleton IR, Westgate K, Samuels TA, Rose AMC, et al. A cross-sectional study of physical activity and sedentary behaviours in a Caribbean population: combining objective and questionnaire data to guide future interventions. BMC Public Health [Internet]. 2016;16(1):1036. Available from: 10.1186/s12889-016-3689-2

50. Huygens MWJ, Swinkels ICS, de Jong JD. Self-monitoring of health data by patients with a chronic disease: does disease controllability matter? BMC Fam Pract. 2017;18:40.

